# Neurodevelopment in children born to women exposed to pesticides during pregnancy

**DOI:** 10.1101/2025.05.25.25328321

**Authors:** William Nelson Mwakalasya, Simon Henry Mamuya, Karim Manji, Bente Elisabeth Moen, Aiwerasia Vera Ngowi

## Abstract

With the global rise in pesticide use, particularly in Africa, pregnant women working in horticulture face a significant risk of pesticide exposure, which may adversely affect fetal neurodevelopment. This study aims to investigate the relationship between self-reported maternal pesticide exposure during pregnancy and neurodevelopmental outcomes in their children, aged 4 to 6 years.

A cross-sectional design was employed, involving 432 mother-child pairs from three horticulture-intensive regions in Tanzania. Maternal exposure to pesticides was assessed by interviewing the mothers, using self-reported questions about activities during pregnancy, including pesticide spraying, weeding, and handling contaminated clothing. Children’s neurodevelopment was evaluated using the International Development and Early Learning Assessment (IDELA), which measures domains such as motor skills, literacy, numeracy, social-emotional development, and executive function.

The children of mothers reporting engaging in pesticide spraying during pregnancy scored significantly lower in social-emotional (β = −6.813, 95% CI: −11.53 to −2.096, p = 0.005) and executive function (β = −9.317, 95% CI: −16.007 to - 2.627, p = 0.006) domains, using linear regression analyses. Children exhibited the highest performance scores in fine and gross motor skills (mean = 62.11±19.3), while the lowest mean scores were in executive function (43.97±24.3). The study also found that older children (aged 6 years) consistently outperformed younger children across all developmental domains.

These findings indicate a relationship between maternal pesticide exposure during pregnancy and adverse neurodevelopment outcomes of their children. However, as the pesticide exposure is based on self-reports, the results should be interpreted with caution. There is a clear need for comprehensive risk assessments that include objective pesticide exposure measurements, particularly in horticulture settings where women of reproductive age constitute a significant portion of the workforce.

## Introduction

Global pesticide use has risen by about 80% in the last two decades [1], with Africa experiencing a staggering 175% increase, largely due to the growing population that put pressure on the agriculture sector to ensure sufficient yields in crop production [2]. However, pesticide use in Africa is associated with severe health consequences for agricultural workers and their families, including increased healthcare costs and long-term adverse effects [3,4]. Despite these risks, most African countries lack the resources to effectively monitor or mitigate the dangers of pesticide exposure, leaving vulnerable populations at risk.

Women are integral to agriculture, especially in low- and middle-income countries. In sub-Saharan Africa, they make up approximately 55% of the agricultural workforce, with Tanzania having the highest proportion at 81% [5]. Within the horticultural sector, women face an increased risk of pesticide exposure through activities such as planting, weeding, harvesting, and pesticide application [6]. Compared to men, women are often more vulnerable to pesticide exposure because they receive less training, and their work is generally perceived as less hazardous, resulting in inadequate protection measures [7]. Exposure pathways include direct contact with pesticides during pesticide mixing and application, as well as indirect exposure from residues on crops, soil and contaminated environments. In addition to working in agriculture, women often engage in domestic tasks that further increase their pesticide exposure. Activities such as washing their husband’s pesticide-contaminated clothing, cleaning spray equipment, and handling other household chores may expose them to additional risks, especially in settings where indoor pesticide spraying is common.

Horticultural work during pregnancy is common in many settings [8,9], and such activities may expose pregnant women to pesticides. Of particular concern is the potential for several of these pesticides to cross the placental barrier, leading to fetal exposure and adverse pregnancy or developmental outcomes [10,11]. The fetus is especially vulnerable to pesticide toxicity due to underdeveloped detoxification mechanisms; for instance, paraoxonase 1 (PON1), an enzyme critical for metabolizing organophosphate pesticides, does not reach adult levels until approximately two years of age [12]. In

Tanzania, several pesticides with documented reproductive, developmental, and neurotoxic effects were reported to be in use in 2020, for instance 2,4-D amine salts, chlorpyrifos, pirimicarb, permethrin, and thiacloprid [13]. Studies have shown relationship between prenatal exposure to hazardous pesticides and neurodevelopmental impairments in children [14,15], mediated through mechanisms such as acetylcholinesterase (AChE) inhibition [16], transcriptomic disruptions in the brain [17], and epigenetic modifications. However, some studies report no significant association between pesticide exposure and neurodevelopment, including research on organophosphate-exposed 2-year-old Chinese children and chlorpyrifos-exposed 36-month-old children of African American and Dominican women in the USA [18,19]. Given these inconsistencies in the literature, further investigation is warranted to clarify the potential risks of prenatal pesticide exposure on neurodevelopmental outcomes.

Pesticide exposure assessment studies vary, as some focus on exposure during the first and second trimesters, while others assess exposure during later stages of pregnancy. Also, neurodevelopmental outcomes have been evaluated using diverse assessment tools across different age groups [20], yet children aged 4 to 6 years have rarely been studied.

The present study focusses on pesticide exposure during the first trimester, as this might be the most vulnerable phase of the pregnancy, related to the development of the fetus. The pesticide exposure information will be obtained by self-reported maternal horticultural activities involving contact with pesticides. Child neurodevelopment was evaluated using the International Development and Early Learning Assessment (IDELA), a standardized tool that measures early learning and developmental milestones, including language and literacy, motor skills, socio-emotional development, numeracy, problem-solving, and learning approaches through interactive play. This study aims to investigate the relationship between self-reported maternal pesticide exposure during pregnancy and neurodevelopmental test outcomes in their children, aged 4 to 6 years. The findings will be useful for development of risk assessments and guidelines related to pesticide exposure during pregnancy.

## Materials and Methods

### Study design and population

This was a cross-sectional analytical study conducted in three regions (A, B and C) of Tanzania. The study was conducted among children born of women working in small-scale horticulture farms who had been working with pesticides before and during conception. Study participants were recruited from two horticulture-intensive wards per district. This is a part of a larger study, and a detailed study plan is described elsewhere [8]. A systematic sampling technique was employed whereby every third household was selected and included in the study. A total of 432 mother-child pairs were obtained, distributed proportionally in the districts based on horticultural land size. Data was collected by five trained research assistants using a digital questionnaire on tablets, with data uploaded daily to Kobo Toolbox servers. Information about the children was collected from their mothers but in order to obtain children’s nutrition status, weight and height of a child was measured. The data collection started 1^st^ November 2022 and ended 15^th^ April 2023.

### Maternal pesticides exposure assessment

Self-reported pesticides exposure during pregnancy was done using a semi-structured closed-ended questionnaire. The questionnaire was used in order to gain information about work and potential exposure to pesticides, as this information is not routinely collected in the farm or health facility records. The questionnaire was divided into two sections. The first section gathered background details such as age, alcohol consumption, cigarette smoking, proximity to the farm, years of experience in horticulture, years lived in the horticulture community, pregnancy-related hospital visits and delivery complications. The second section examined activities performed by women that could contribute to pesticide exposure during pregnancy. Participants were asked to recall whether, during the pregnancy of the child under study, they engaged in any of the following activities: mixing or diluting pesticides, spraying pesticides, weeding or harvesting within 24 hours after pesticide application, washing spray equipment, laundering pesticide-contaminated clothing, or consuming vegetables or fruits within 24 hours of pesticide application.

### Neurodevelopmental assessment

The International Development and Early Learning Assessment (IDELA), developed by Save the Children, was used to measure children’s learning and development across various domains, including motor, literacy, numeracy, and social-emotional development (Table 1). IDELA is a child centered tool that comprises 24 core items designed to directly assess key developmental skills and early learning in preschool children aged 3.5 to 6.5 years. The IDELA tool has already been utilized in over 70 countries [21] demonstrating its adaptability to diverse cultural contexts and its strong reliability and validity. For this study, the original English version of IDELA was translated into Kiswahili, the native language of both the researchers and the study participants. The assessment was conducted at home by trained research assistants, who first obtained the child’s assent before securing verbal consent from the mother or caregiver. The mother/caregiver remained present throughout the assessment to ensure the child felt safe and comfortable.

**Table 1:**
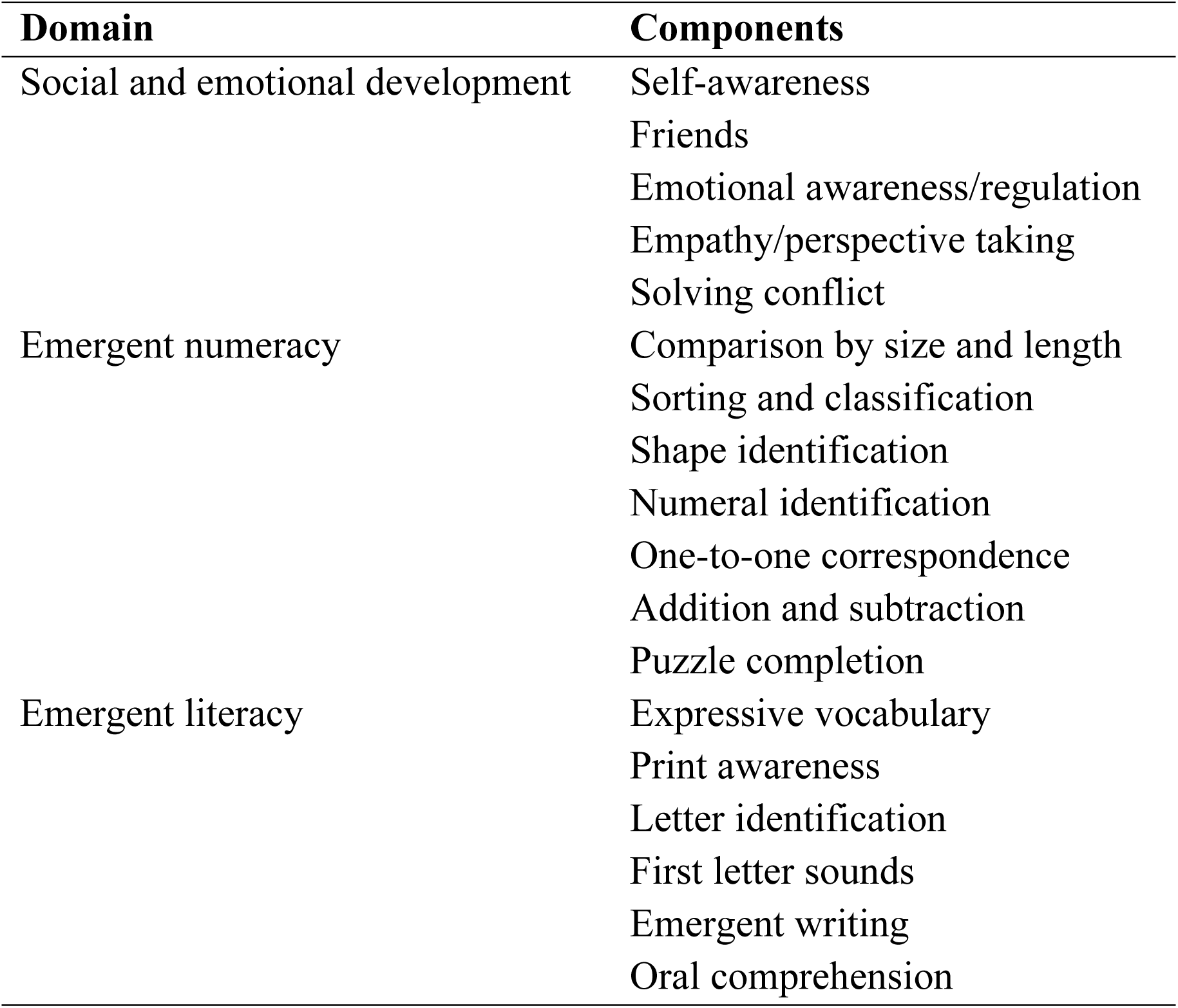
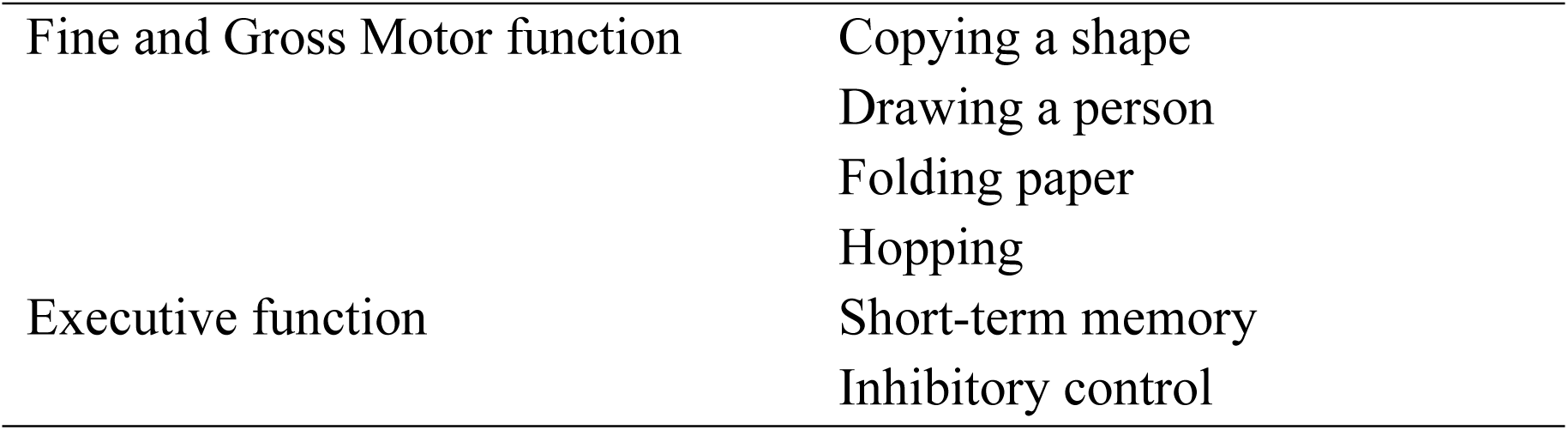
Components of the IDELA domains.

### Statistical analysis

Descriptive statistics were used to summarize the independent variables, which included sociodemographic factors and pesticide exposure activities reported by women during pregnancy. The dependent variable was the IDELA scores, representing the points accumulated by children across five neurodevelopmental domains: social-emotional, emergent numeracy, emergent literacy, fine and gross motor skills, and executive function. These scores were converted into percentages, and total scores were calculated by summing points from all domains. Higher domain mean scores indicated better performance.

The Shapiro-Wilk test for normality revealed that the neurodevelopmental scores were not normally distributed (p<0.05). Consequently, a nonparametric Independent-Samples Mann-Whitney U Test was used for a univariate analysis to examine the relationship between the independent variables and the dependent variable mean scores. The significance level was set to p<0.01, due to the large number of analyses. Additionally, a multiple linear regression analysis was conducted to assess the relationship between the independent and dependent variables (domain specific and total neurodevelopment score). Independent variables were the pesticide exposure-related activities and maternal and child social demographic characteristics. The dependent variable was each of the neurodevelopment scores; they were analyzed separately. All statistical analyses were performed using the Statistical Package for the Social Sciences (SPSS), version 27.

### Ethics approval and consent to participate

The study was approved by the Institutional Review Board of Muhimbili University of Health and Allied Sciences (MUHAS) (MUHAS-REC-08-2022-1332) and the Regional Committee for Research Ethics of Southeast Norway (535644/2023). District executive directors granted permission to access wards, villages, and households, issuing letters to inform Ward Executive Officers (WEOs), who in turn notified hamlet leaders by phone. During household or farm visits, research assistants explained the study’s purpose, and participants provided verbal consent before the interview. The consent was documented in written by a researcher, and witnessed by a supervisor with writing skills at the workplace. During neurodevelopmental assessment a child had to assent before obtaining a verbal consent from a mother/caregiver.

## Results

### Social-demographic characteristics of study participants

In this study, a total of 432 mother-child pairs out of 436 reached (99% response rate) were included in the analysis. Four women declined to participate, citing dissatisfaction with the perceived benefits of the study. Most mothers were aged 31-40 years (55.1%) and over half (55.3%) had six to ten years of experience in horticulture, but most (39.8%) have been living in the area for more than 20 years. Delivery complications were reported by only few (3.7%) women. Children were dominated by those aged 6 years (43.5%) and sex distribution was almost equal with girls slightly outnumbering boys (53% vs. 47%). Re-entry in the farm for weeding within 24 hours after spray (57.2%) and washing clothes used for pesticide spray (51.6%) are the activities most reported by women (**Table 2**).

**Table 2:** Social demographic characteristics of 432 mother and child pairs.

| Variables | Categories | Frequency (%) |
| --- | --- | --- |
| **Maternal characteristics** |  |  |
| Study areas | A | 80 (18.5) |
|  | B | 258 (59.7) |
|  | C | 94 (21.8) |
| Mother's age (years) | Under 30 | 141 (32.6) |
|  | 31 - 40 | 238 (55.1) |
|  | Above 40 | 53 (12.3) |
| Years in horticulture | Under 5 | 122 (28.2) |
|  | 6 to 10 | 239 (55.3) |
|  | Above 10 | 71 (16.4) |
| Delivery complications | No | 416 (96.3) |
|  | Yes | 16 (3.7) |
| Years lived in the area | Under 10 | 138 (31.9) |
|  | 11 to 20 | 122 (28.2) |
|  | Above 20 | 172 (39.8) |
| **Children characteristics** |  |  |
| Age (years) | 4 | 104 (24.1) |
|  | 5 | 140 (32.4) |
|  | 6 | 188 (43.5) |
| Sex | Boys | 203 (47.0) |
|  | Girls | 229 (53.0) |
| Nutrition status | Normal ($-1 \leq \text{WHZ} \leq +1$) | 330 (77.1) |
| | Wasted ($\text{WHZ} < -1$) | 54 (12.6) |
| | Overweight ($\text{WHZ} > +1$) | 44 (10.3) |
| **Work with possible pesticides exposure practiced during pregnancy** |  |  |
| Mixing pesticides | No | 398 (92.1) |
|  | Yes | 34 (7.9) |
| Spraying pesticides | No | 269 (62.3) |
|  | Yes | 163 (37.7) |
| Weeding within 24 hours after spray | No | 185 (42.8) |
|  | Yes | 247 (57.2) |
| Harvesting within 24 hours after spray | No | 335 (77.5) |
|  | Yes | 97 (22.5) |
| Washed clothes used in pesticides work | No | 209 (48.4) |
|  | Yes | 223 (51.6) |
| Washing spray equipment after use | No | 302 (69.9) |
|  | Yes | 130 (30.1) |
| Eating crops within 24 hours after spray | No | 235 (54.4) |
|  | Yes | 197 (45.6) |

### Children’s neurodevelopment performance

The mean IDELA scores by domain ranged between 43.59% to 62.11% with an overall mean of 50.69%. Fine and gross motor domain had the highest mean score of 62.11±19.3 followed by emergent numeracy with a mean score of 57.04±23.1 and the lowest mean scores was achieved in emergency literacy (43.59±20.7) and executive function (43.97±24.3) domains (Fig. 1).

**Figure 1:**
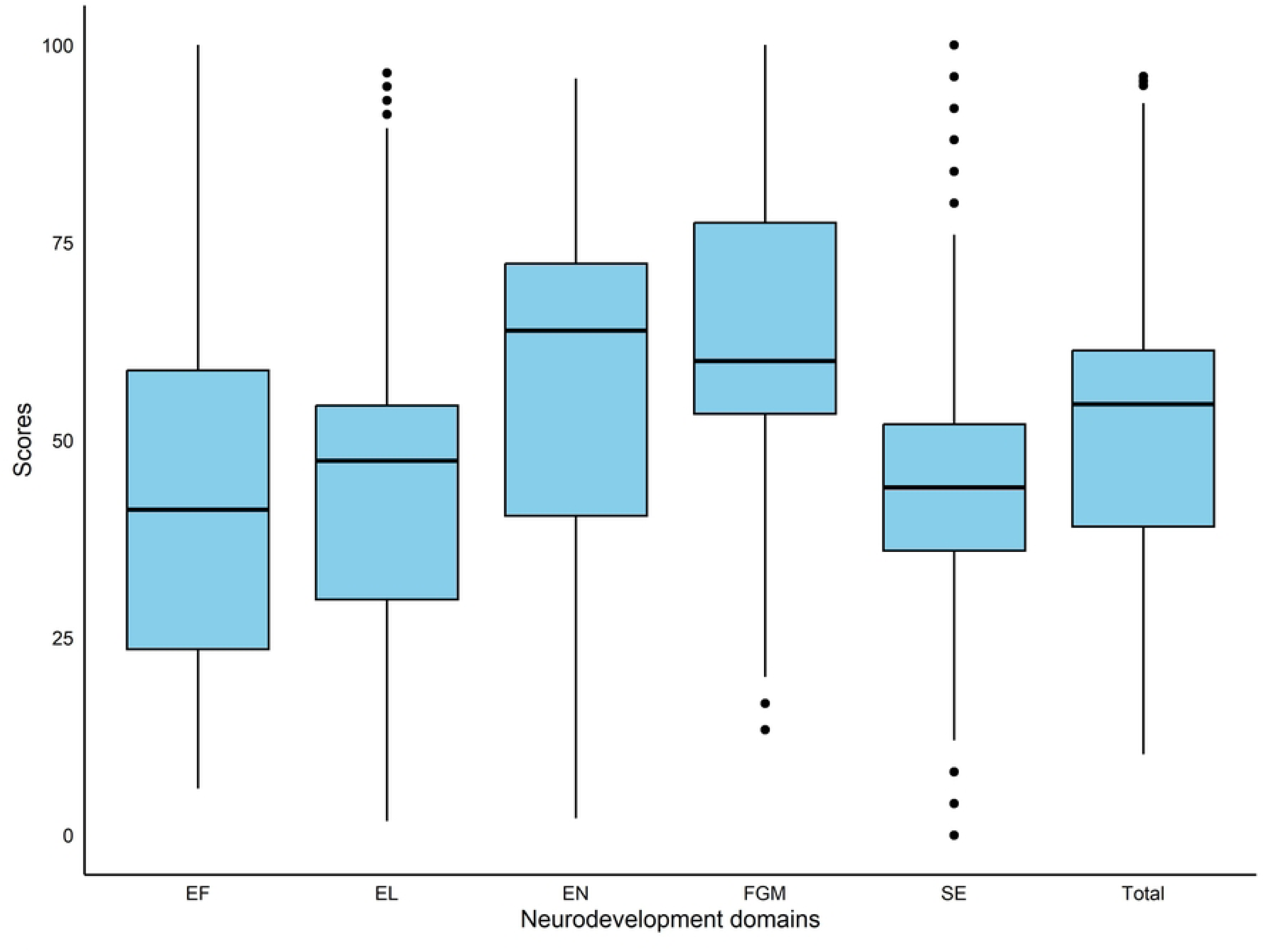
A boxplot showing the distribution of mean neurodevelopment scores for each domain. SE – Social emotional; EN – Emotional Numeracy; EL – Emergent Literacy; FGM – Fine and Gross Motor; EF – Executive Function Independent-Samples Mann-Whitney U Test showed that child’s age was significantly associated with the neurodevelopment scores across all domains (p≤0.01) (Table 3). Older children (6 years) consistently exhibited higher mean scores across all domains compared to younger children, with six-year-olds displaying the highest scores. Other sociodemographic factors, such as study area, mother’s age, child’s sex, years in horticulture, years lived in the area, and delivery complications, did not show statistically significant differences with neurodevelopment scores.

**Table 3:**
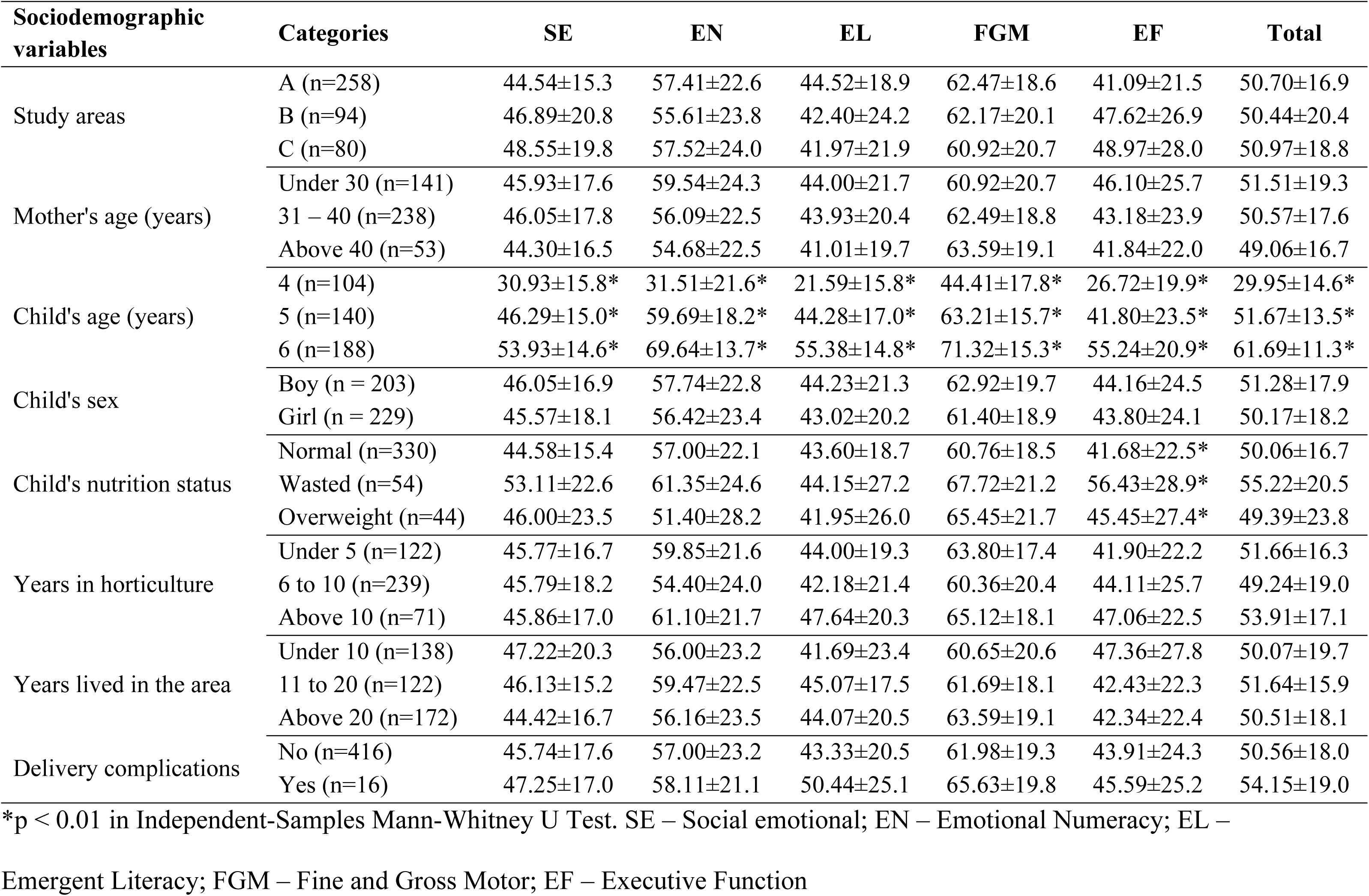
The association between maternal and child sociodemographic variables and mean neurodevelopment scores in 432 children aged 4 – 6 years.

### The association between maternal pesticide exposure and **their c**hildren’s neurodevelopment (performance)

Women who reported to engage in pesticides spray activity during pregnancy had children with significantly lower mean emergent numeracy (53.88±20.1) and lower fine and gross motor domain scores (58.77±17.8) compared to those who reported to not spray during pregnancy (Mann-Whitney U-test, p≤0.01) (Table 4). Similar outcomes were observed in children of women who reported washing clothes used for pesticides spray/handling activities. Mean emergent numeracy domain scores were also significantly lower (54.88±22.6) in children of mothers who reported to engage in weeding activities within 24 hours after pesticide spray during pregnancy, compared to those who reported not to (59.93±23.4). Other horticulture practices like mixing pesticides, washing equipment used for pesticides work and eating crops sprayed with pesticides within 24 hours did not show any statistically significant relationship with the neurodevelopment scores.

**Table 4:** The association between maternal pesticide exposure variables and mean neurodevelopment scores in children aged 4 – 6 years.

| Pesticides exposure activities practiced during pregnancy | Categories | SE | EN | EL | FGM | EF | Total |
| --- | --- | --- | --- | --- | --- | --- | --- |
| Mixing pesticides | No (n=398) | 45.82±17.7 | 57.30±23.4 | 43.45±20.7 | 62.55±19.3 | 43.91±24.2 | 50.79±18.2 |
|  | Yes (n=34) | 45.53±15.3 | 54.01±19.3 | 45.20±20.2 | 57.06±18.3 | 44.64±25.4 | 49.57±15.6 |
| Spraying pesticides | No (n=269) | 46.30±18.8 | 58.96±24.6* | 44.03±21.6 | 64.14±19.9* | 44.54±25.4 | 51.82±19.4 |
|  | Yes (n=163) | 44.96±15.3 | 53.88±20.1* | 42.87±19.2 | 58.77±17.8* | 43.02±22.4 | 48.83±15.4 |
| Weeding within 24 hours after spray | No (n=185) | 45.02±16.2 | 59.93±23.4* | 45.04±19.5 | 64.76±19.0 | 42.58±22.8 | 52.13±18.1 |
|  | Yes (n=247) | 46.38±18.5 | 54.88±22.6* | 42.51±21.5 | 60.13±19.3 | 45.01±25.3 | 49.61±18.0 |
| Harvesting within 24 hours after spray | No (n=335) | 45.98±17.1 | 57.65±23.1 | 44.18±20.4 | 63.00±19.3 | 43.76±23.7 | 51.20±17.8 |
|  | Yes (n=97) | 45.15±19.0 | 54.95±23.2 | 41.54±21.5 | 59.07±19.1 | 44.69±26.3 | 48.93±18.6 |
| Washed clothes used in pesticides work | No (n=209) | 44.86±16.8 | 59.03±24.1* | 45.03±20.1 | 64.32±19.2* | 42.08±22.8 | 51.75±18.2 |
|  | Yes (n=223) | 46.67±18.2 | 55.18±22.0* | 42.25±21.2 | 60.04±19.2* | 45.74±25.4 | 49.70±17.8 |
| Washing spray equipment after use | No (n=302) | 45.14±16.7 | 57.46±23.1 | 44.33±20.0 | 62.41±19.4 | 42.46±22.6 | 50.85±17.7 |
|  | Yes (n=130) | 47.32±19.3 | 56.07±23.0 | 41.88±22.1 | 61.44±19.1 | 47.47±27.5 | 50.31±18.7 |
| Eating crops within 24 hours after spray | No (n=235) | 45.09±15.5 | 58.76±22.0 | 44.68±19.1 | 63.13±18.9 | 42.53±22.0 | 51.44±16.8 |
|  | Yes (n=197) | 46.64±19.7 | 55.00±24.3 | 42.29±22.4 | 60.90±19.8 | 45.69±26.6 | 49.80±19.4 |

\*p < 0.01 in Independent-Samples Mann-Whitney U Test. SE – Social and emotional development; EN – Emergent Numeracy; EL – Emergent Literacy; FGM – Fine and Gross Motor function; EF – Executive Function

A multiple linear regression, with all independent variables included in the model simultaneously, was run to predict neurodevelopment scores from reported pesticide exposure activities using the child’s age as a covariate (Figure 2). The analysis showed a borderline relationship between engaging in spray work during pregnancy and low total test scores (β = −4.127, 95% CI: −8.256 to 0.002, p = 0.05), while no other statistical relationships were found. It was also shown that spraying pesticides during pregnancy was significantly associated with lower social-emotional scores (β = −6.813, 95% CI: −11.53 to −2.096, p = 0.005) and executive function scores (β = −9.317, 95% CI: −16.007 to −2.627, p = 0.006). On the other hand, children of women who reported doing weeding within 24 hours after spraying showed a significant positive association with social-emotional scores (β = 4.774, 95% CI: 0.174 to 9.373, p = 0.042). Emergency numeracy, emergency literacy and fine and gross motor scores were not associated to any of the variables in the model.

**Figure 2:**
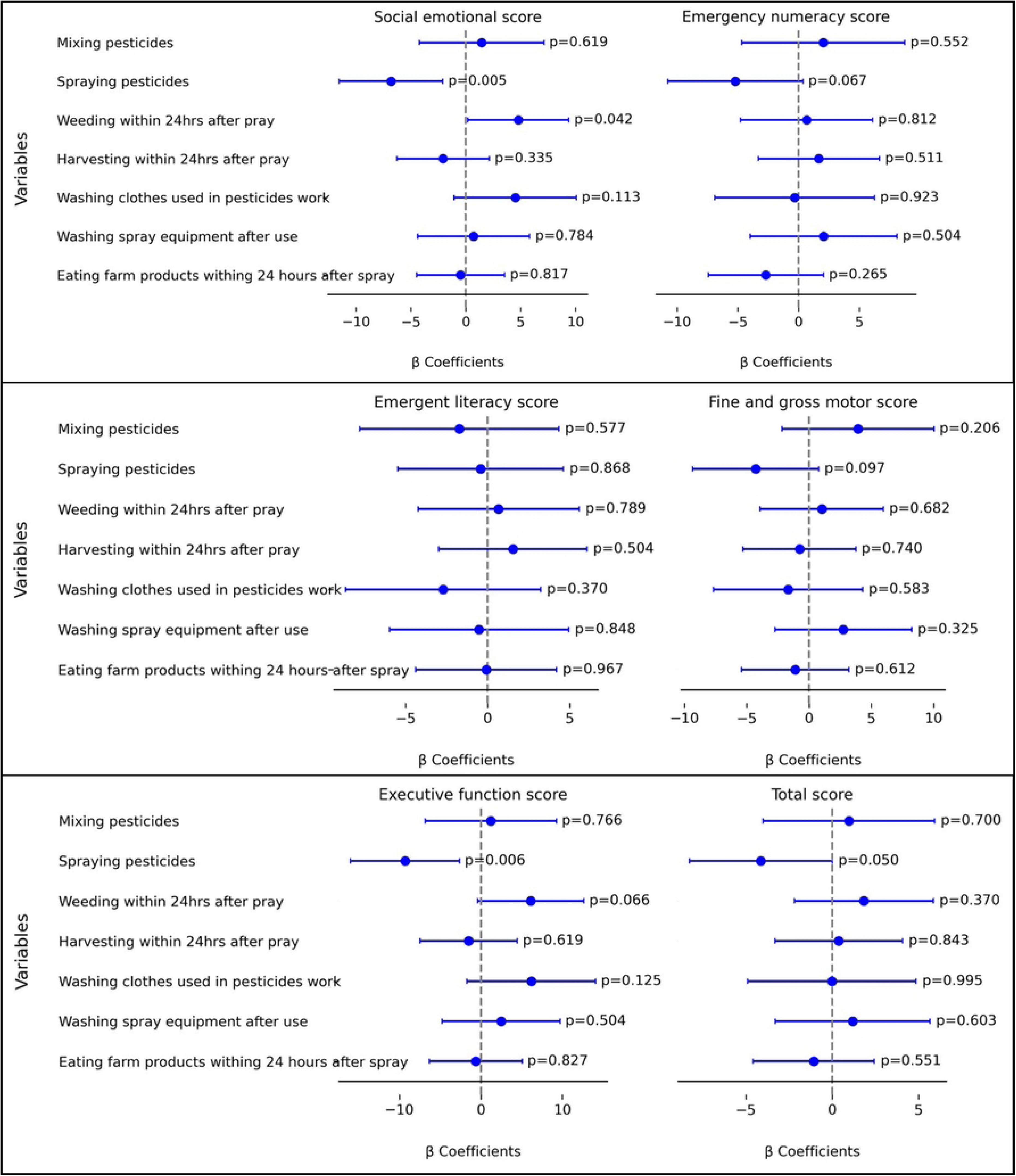
The associations between 7 maternal self-reported pesticide exposure situations during pregnancy and their children’s performance on the neurodevelopment tests IDELA domains at the of 4 to 6 years in Tanzania. Adjusted β coefficients and 95% confidence interval are shown as results from multiple linear regression analysis, adjusted for the child’s age.

## Discussion

Our results show that self-reported maternal exposure to pesticides through direct spraying during pregnancy was associated with lower scores in social-emotional and executive function domains among children aged 4 to 6 years. We also found an association between the social-emotional scores among the children and of the engagement in weeding among their mothers during pregnancy. Additionally, direct spraying of pesticides showed a borderline association with reduced overall neurodevelopmental scores.

These results are consistent with a previous study conducted among children of smallholder tomato farmers in southern Tanzania, which reported delayed neurodevelopment in children associated with maternal engagement in agricultural work [22]. However, our present study provides more granular insight by identifying direct pesticide spraying as a significant contributor to maternal exposure. This distinction helps bridge an important gap in understanding how specific horticultural practices during pregnancy relate to adverse neurodevelopmental outcomes in children.

Adverse effects of pesticide exposure on social-emotional development have also been shown in several other studies. In Costa Rica, a prospective neurodevelopmental assessment of 355 one-year-old infants, born to mothers living within 5 km of banana plantations with frequent aerial mancozeb (a fungicide) spraying, found that higher prenatal urinary ethylenethiourea (a mancozeb metabolite) concentrations were associated with lower social-emotional scores, particularly among girls [23]. Similarly, a study analyzing 618 urine samples from women exposed to insecticides used in indoor residual spraying for malaria control in Limpopo, South Africa, revealed that maternal urinary metabolites of certain pyrethroids, specifically cis–(2,2-dichlorovinyl)-2,2 dimethylcyclopropane-1-carboxylicacid (DCCA), *trans*-DCCA, and 3-phenoxybenzoic acid (3PBA), were associated with lower social-emotional scores in one-year-old infants [24]. Postnatal exposure to organochlorine pesticides through breastfeeding has also been linked to impaired social-emotional development [25], though prenatal exposure did not show the same association [24]. Supporting this, a study of 55 Taiwanese infants aged 8–12 months found that higher breast milk levels of some organochlorines were significantly associated with poorer social-emotional performance [25]. Together, these results suggest that both prenatal and postnatal pesticide exposure may adversely affect early social-emotional development.

In accordance with the present results on the association between pesticides exposure and executive functioning in children, previous studies have demonstrated similar results. In a U.S birth cohort involving 162 mother-child pairs found that prenatal exposure to pyrethroids, as indicated by its urinary metabolites 3-PBA and cis-DCCA, was associated with poorer performance in executive function in children aged 4 to 9 years [26]. Similarly, the CHAMACOS cohort study involving 363 mother-child pairs reported that higher gestational concentrations of organophosphate metabolites, as measured by maternal urine levels of dialkyl phosphate, were associated with significantly lower executive functioning in children aged between 7 and 12 years [27]. These findings in previous studies and the present one, suggest that early-life exposures to neurotoxic pesticides may contribute to impairments in core executive function domains such as short-term memory and inhibitory control.

There is also some inconsistence in the relationship between prenatal pesticide exposure and child neurodevelopment. For instance, in a Canadian cohort, first trimester maternal urinary glyphosate levels were not significantly associated with child neurodevelopment scores at 3 to 4 years of age [28]. Similarly, a study in Mexico City found that third-trimester levels of 3-PBA was not associated with changes in mental or motor development scores by 36 months [29]. In a Colombian cohort, where 13% of mothers reported spraying pesticides in the farm during pregnancy, the children had slightly lower but not statistically significant neurodevelopmental scores [30]. These evidences suggest that while some pesticide exposures show impact in children’s early neurodevelopment, there are still inconsistencies among the studies which may be attributed to differences in exposure timing, type of pesticide studied, and how exposure and outcomes were measured. More research work is needed.

No significant associations were found in the present study between pesticide exposure from activities such as mixing pesticides, washing spray equipment, harvesting within 24 hours after pesticide spray or consuming crops within 24 hours of spraying. This may reflect differences in exposure intensity or variability in protective behaviors. For example, pesticide mixing may result in relatively lower exposure since it is less commonly performed by women, and when they do engage in it, they tend to use protective gear more consistently compared to when spraying [31]. Regarding pesticide ingestion, a quasi-experimental study of rural school children in Chile found that fruit consumption was associated with elevated concentrations of only one out of ten studied organophosphate metabolites [32]. This suggests that ingestion may not be a primary exposure route. However, findings from South Africa indicate that school children who consumed fruits directly from vineyards or orchards exhibited lower motor screening speed and visual processing accuracy scores [33].

Harvesting, a task predominantly performed by women [34], did not present a significant exposure risk in this study. In Kenya, individuals involved in harvesting on horticultural farms reported a higher frequency of pesticide-related symptoms [35]. Thus, pesticide exposure through harvesting exists and the study underlines that harvesting is mostly performed by women. In our study, children of mothers who engaged in harvesting had significantly lower fine and gross motor scores, though this association was not observed in the adjusted statistical models.

Children in this study population demonstrated higher neurodevelopmental scores (mean IDELA score: 51%) compared to those from other low-resource setting countries. The overall mean IDELA scores in this study were slightly higher than those reported for children aged 3–6 years in Cape Town, South Africa (mean score: 48%) [36] . The scores in the present study were clearly higher than among children aged 3–5 years, in a study from 29 villages in Cambodia (mean score: 43%) [37]. This difference may be partly attributed to the age distribution of the study populations, which was 4 to 6 years in the current study, as all studies noted an upward trend in scores with increasing age. However, the scores in our present study were lower than those of preschool-aged children from low socioeconomic backgrounds in São Paulo, Brazil, who achieved an average IDELA score of 76.4% [38]. It is difficult to know why these differences are seen, but it might be related to different cultures and early education of the children [39].

This study demonstrates several methodological strengths; it is the first study to employ IDELA, a standardized and validated tool for objective evaluation of neurodevelopmental outcomes in children aged 3.5 to 6.5 years in low- and middle-income countries. By using IDELA, the study ensures precise and consistent assessments across critical developmental domains, including cognitive, motor and social emotional skills. Previous pesticide exposure studies used the Bayley Scales of Infant Development [40,41] and the Cambridge Automated NeuroPsychological Battery [33], yielding consistent findings. The children in our study had different age, and this may have influenced the results, as older children perform better than younger ones. This was controlled in the study by using statistical analyses to adjust for the age of the children. This adjustment was pivotal in ensuring that the observed relationships represent a more precise attribution of observed neurodevelopmental outcomes to pesticide exposure rather than extraneous factors.

However, the study has several limitations. The reliance on self-reported data to assess pesticide exposure introduces potential recall bias, a common issue in similar research [42]. Such bias may compromise the validity of exposure estimates. Previous studies indicate that the accuracy of self-reported pesticide exposure depends on the timing and specificity of recall. When collected shortly after exposure or focused on general exposure information (e.g., hygiene practices), self-reports can yield relatively accurate predictions of outcomes like neurodevelopmental scores [43,44]. However, in this study, women were asked to recall farm activities from four to six years prior, which may have affected precision. Also, the cross-sectional design poses inherent limitations, though these were mitigated through statistical methods examining associations between variables. While the study conducted multiple statistical tests, the potential for bias due to this was reduced by adopting a stricter significance threshold (p < 0.01).

In conclusion, our findings suggest that maternal involvement in pesticide-related activities, specifically pesticide spraying during pregnancy is associated with lower neurodevelopment scores, particularly in the social-emotional, executive function, and overall domains of their children. As the pesticide exposure is based on self-reports, these results should be interpreted with caution. Risk assessments, including pesticide exposure measurements, are clearly needed in horticulture, especially where the workers are women of reproductive age.

## Data Availability

Data cannot be shared due to restrictions from the Ethical committees - it is to protect the participants.

## Acknowledgments

We sincerely appreciate the support of the government authorities in the study areas during data collection. Most importantly, we are deeply grateful to all the women who participated in this study—their experiences and insights were invaluable in making this research possible.

